# Cognitive functioning and lifetime Major Depressive Disorder in UK Biobank

**DOI:** 10.1101/19006031

**Authors:** L De Nooij, MA Harris, MJ Adams, T-K Clarke, X Shen, SR Cox, AM McIntosh, HC Whalley

**Affiliations:** Division of Psychiatry, University of Edinburgh, UK; Centre for Cognitive Ageing and Cognitive Epidemiology, University of Edinburgh, UK; Department of Psychology, University of Edinburgh, UK

**Keywords:** cognition, depression, psychosocial functioning, medication, UK Biobank

## Abstract

**Background:** Cognitive impairment associated with lifetime Major Depressive Disorder (MDD) is well-supported by meta-analytic studies, but population-based estimates remain scarce. Previous UK Biobank studies have only shown limited evidence of cognitive differences related to probable MDD. Using updated cognitive and clinical assessments in UK Biobank, this study investigated population-level differences in cognitive functioning associated with lifetime MDD.

**Methods:** Associations between lifetime MDD and cognition (performance on six tasks and general cognitive functioning (*g-*factor)) were investigated in UK Biobank (*N*-range 7,457-14,836, age 45-81 years, 52% female), adjusting for demographics, education and lifestyle. Lifetime MDD classifications were based on the Composite International Diagnostic Interview. Within the lifetime MDD group, we additionally investigated relationships between cognition and (i) recurrence, (ii) current symptoms, (iii) severity of psychosocial impairment (while symptomatic), and (iv) concurrent psychotropic medication use.

**Results:** Lifetime MDD was robustly associated with a lower *g*-factor (*β* = −0.10, *P*_*FDR*_ = 4.7×10^−5^), with impairments in attention, processing speed and executive functioning (*β* ≥ 0.06). Clinical characteristics revealed differential profiles of cognitive impairment among case individuals; those who reported severe psychosocial impairment and use of psychotropic medication performed worse on cognitive tests. Severe psychosocial impairment and reasoning showed the strongest association (*β* = −0.18, *P*_*FDR*_ = 7.5×10^−5^).

**Conclusions:** Findings describe small but robust associations between lifetime MDD and lower cognitive performance within a population based sample. Overall effects were of modest effect size, suggesting limited clinical relevance. However, deficits within specific cognitive domains were more pronounced in relation to clinical characteristics, particularly severe psychosocial impairment.

## 1. Introduction

Major Depressive Disorder (MDD) is a highly prevalent condition, affecting around one in five people in their lifetime globally (1–3). Previous clinical research has shown that individuals with MDD show cognitive deficits, particularly in executive functioning, working memory, attention, and processing speed (4–7), as well as affect-related cognitive biases, including negative information biases for perception, attention, and memory (8,9). Additionally, there is evidence that residual cognitive deficits are present in remitted cases (4,10–12). A recent systematic review and meta-analysis that investigated studies including individuals remitted from MDD revealed significant small to moderate deficits in the same domains of cognitive functioning, and showed worse cognitive functioning associated with recurrent episodes (12).

Meta-analyses are invaluable for investigating patterns of results from smaller studies that are heterogenous in nature. One disadvantage however is that included studies show differential procedures of participant inclusion and recruitment, of which some will be more focused on certain clinical populations (e.g. allowing bipolarity or including participants after antidepressant treatment). As a result, meta-analytically derived effect sizes may overestimate cognitive impairment related to MDD compared to the general population. Understanding the degree of impairment associated with lifetime MDD (current or past diagnosis) in the general population thus requires further investigation within a sufficiently powered community-based sample. This summarises the added value of population-based studies which are not biased by specific clinical characteristics and that are sufficiently powered to detect potentially modest effect sizes.

UK Biobank is a large-scale adult population-based study that is well-suited to investigate population-level cognitive differences related to lifetime MDD. Previous population-based studies that investigated baseline assessments of the UK Biobank cohort study indicated modestly decreased visuospatial memory performance in participants with lifetime MDD (13), but showed comparable cognitive performance to controls for other cognitive measures of reasoning, reaction time and memory (13,14). Robust evidence from population-based studies is currently limited, so that the degree and patterns of cognitive impairment in community-based individuals with lifetime MDD remain uncertain.

Of note, UK Biobank follow-up assessments were recently extended to include updated cognitive measures with improved reliability (15,16). These tests also cover a broader range of cognitive domains previously implicated in MDD and show sufficient performance variability within the healthy population. Furthermore, previous mood disorder groupings relied on a combination of self-report and relatively unstructured questionnaire items, whereas the more recent lifetime MDD assessments were based on a structured diagnostic interview questionnaire, i.e. the Composite International Diagnostic Interview (CIDI-SF) (17). In the current study we used the opportunity provided by these updated assessments to further investigate patterns of cognition functioning in lifetime MDD within the large population-based UK Biobank sample.

Furthermore, the novelty and clinical relevance of this study was increased by additionally conducting a population-based investigation of clinical characteristics associated with cognitive functioning. The first clinical variable of interest was recurrence of depressive episodes, given the usually highly recurrent nature of MDD and the ongoing discussions with regard to ‘scar theories’ (which propose that disease-related psychological or biological changes may result in a predisposition to future depressive episodes) (18). Although earlier research did not consistently report recurrence of MDD being associated with lower cognitive functioning (10,19), recent evidence suggests that cognitive deficits do accumulate with repeated episodes (12). Furthermore, although no optimal measure of MDD remission was available from UK Biobank assessments, we investigated associations with the putative presence of current depressive symptoms. There is a general consensus that low mood can negatively affect cognitive performance, and more pronounced cognitive impairment for symptomatic individuals is supported by previous meta-analytic studies (6,10,20). However, another meta-analysis showed similar effect sizes for the current and remitted state (4), and associations with depressive symptoms may be attenuated within the general population (21). Thirdly, we addressed associations between psychosocial functioning during the depressive episode and subsequent cognitive deficits. Previous research suggests that cognitive deficits contribute to impaired psychosocial functioning (22–24), which impacts on quality of life. Furthermore, cognitive deficits were found to mediate of decreased work performance, contributing to the overall cost attributable to MDD (25,26). The fourth clinical variable of interest was use of psychotropic medication concurrent with cognitive assessment.

Pharmacological treatment is expected to have a complex influence on cognitive functioning, although reliable evidence is limited (27). Meta-analyses suggest that some antidepressants may improve cognition (28,29), but these results appear to be specific to particular treatments (30–32), while for other treatments, medication side-effects may potentially affect cognitive performance negatively (6,14,33). Given the variety of psychotropic medications, we did not aim to investigate their differential effects, but rather investigated the overall association between psychotropic medication and cognitive functioning within the general population.

In summary, the aim of the current study was to investigate cognitive functioning in the context of lifetime MDD within the general population, as conducted within the large-scaled population-based UK Biobank sample. Robustness of associations was tested via inclusion of different combinations of demographic, education and lifestyle (smoking, alcohol consumption and Body Mass Index [BMI] (34,35)) covariates. We also examined clinical characteristics that are considered relevant to cognitive functioning in the context of lifetime MDD and may therefore influence the association: (i) recurrence of depressive episodes, (ii) current MDD symptomatology, (iii) severe psychosocial impairment (while symptomatic), and (iv) use of psychotropic medication (at time of assessment). These characteristics were hypothesised to distinguish case subgroups with more severe cognitive impairment, or specifically for psychotropic medication, either increased or reduced impairment.

## 2. Methods

### 2.1. Participants

Adults (40 to 69 years) were recruited for participation in UK Biobank (http://www.ukbiobank.ac.uk) between 2006 and 2010 (36). The present study included participants with data for at least one of the neuropsychological test scores from the third UK Biobank assessment (2014 onwards; *N* = 28,480). Participants with self-reported neurological conditions were excluded from all analyses. A *g*-factor, representing general cognitive function, was derived from all remaining participants with complete data for included cognitive test measures (*N* = 13,589). When assessing individual tasks, complete cognitive data was not required. Participants with specific psychiatric disorders or an unclassified/missing lifetime MDD status were excluded from further analyses (Supplementary Materials 1, Figure S1). Following exclusion criteria, *N* = 14,877 individuals (*n*_cases_ = 4,486, *M*_age_ = 63.5, *SD*_age_ = 7.4, age range 45-81 years, 52% female) were included across all analyses, with sample sizes ranging from *N* = 7,457 to *N* = 14,386 per case-control analysis.

### 2.2. Measures and procedure

At the third assessment, participants visited the assessment centre where they provided demographic and health information in response to a series of touchscreen questions and completed computerised cognitive tests on a touchscreen. Medical history and current medication use were assessed during interviews led by health professionals. Furthermore, participants completed a web-based mental health questionnaire (MHQ) at home (37). Most participants (all those with complete cognitive data) completed the MHQ between 2.2 years before and 0.4 years after their assessment centre visit. UK Biobank received research ethics committee (REC) approval for the centre assessments (ref 11/NW/0382), later amended to also cover the MHQ, and all participants provided informed consent before assessment (http://biobank.ctsu.ox.ac.uk/crystal/field.cgi?id=200).

#### 2.2.1. Classification of lifetime Major Depressive Disorder

Case-control classification was based on the short form of the Composite International Diagnostic Interview (CIDI-SF) (17), administered as part of the MHQ (37). This included two binary questions regarding (i) ever having experienced depressive feelings or (ii) loss of interest, for a period of two or more weeks. Participants who responded ‘Yes’ to either question then answered a question about the lifetime number of depressive episodes, followed by six binary questions on experience of other DSM-IV MDD symptoms during their worst episode, including: (iii) feelings of worthlessness, (iv) tiredness, (v) difficulty concentrating, (vi) suicidal thoughts, (vii) changes in sleeping pattern, and (viii) changes in weight. Summed responses to all eight depression symptom questions provided a symptom score (range 0-8). Participants were also asked how often they had experienced depressive feelings / loss of interest during their worst episode, how long these feelings lasted, and whether they interfered with their “roles, life or activities” (psychosocial impairment). Participants were classified as lifetime MDD cases if all of the following applied (37):

i. Summed symptom score >= five
ii. At worst, symptoms experienced “almost every day” or “every day”, and lasted “most of the day” or “all day”
iii. Symptoms impaired psychosocial functioning “somewhat” or “a lot”

#### 2.2.2. Clinical characteristics

Within individuals who were classified with lifetime MDD as described above, we further identified four clinical characteristics: (i) recurrent MDD, reflecting more than one depressive episode, (ii) putative current MDD symptoms, as derived from responses on the touchscreen questionnaire (Supplementary Materials 2), (iii) severe psychosocial impairment (at the time of the depressive episode), indicated by the maximum CIDI-SF symptom score of eight in combination with the maximum score for psychosocial impairment (37); and (iv) concurrent use of psychotropic medication, which was derived from reported use of antidepressant (94.6%), antipsychotic (3.9%) or anxiolytic (1.5%) medication as assessed during the nurse-led interview (Supplementary Materials 3, Figure S2).

#### 2.2.3. Cognitive assessment

In the current study we considered cognitive tests from the third UK Biobank assessment. We included data from the following cognitive tests:

i. Digit Symbol Substitution test (DSST)
ii. Trail Making Test, alphanumeric trail (TMT-B)
iii. Numeric Memory (NM)
iv. Matrix pattern completion (Matrix)
v. Verbal Numeric Reasoning (VNR)
vi. Tower rearranging (Tower)

This selection comprises all updated cognitive test data available at time of analysis (DSST, TMT-B, Matrix, Tower), and additionally included two cognitive tests repeated at this assessment that were also administered at earlier UK Biobank assessments (VNR, NM). Further details on each cognitive test can be found in Table 1. Psychometric properties, advantages and limitations of the UK Biobank cognitive tests have been discussed in detail elsewhere (16).

**Table 1.** Descriptions of cognitive tests

| Test | Description | Cognitive domains | Variable | Range |
| --- | --- | --- | --- | --- |
| DSST | Presented with a key grid containing digit-symbol pairs, the participant had one minute to indicate which number corresponded to each of the symbols in the task grid. | Processing speed and attention | Total correct score | 0-89 |
| TMT-B | Digits and letters were scattered around the screen in circles. The participant was asked to click on them sequentially, alternating between digits and letters (1-A-2-B-3-C etc.) | Executive functions (mental flexibility, complex attention) and processing speed | Time to completion (s)<br>*reversed in analyses* | 20-300 |
| NM | Every trial, a sequence of digits was briefly presented on screen, after which the participant was asked to recollect it from memory. If answered correctly, the sequence length increased with one digit. | Working memory and attention | Maximum digits remembered correctly | 2-12 |
| Matrix | Presented with a series of matrix pattern blocks that each missed one element, the participant was asked to select the element that best completed the pattern from a range of choices. | Reasoning ability (abstract) | Total correct score | 0-15 |
| VNR | The participant had two minutes to complete as many multiple choice reasoning questions as possible. | Reasoning ability (verbal, numeric) and processing speed | Total correct score | 0-13 |
| Tower | Presented with a series of illustrations displaying three differently-coloured hoops on three pegs, the participant was asked how many moves it would take to re-arrange the hoops into another specific position. | Executive functions (strategy, planning) and working memory | Total correct score | 0-18 |

DSST, Digit Symbol Substitution Task; Matrix, Matrix pattern completion; NM, Numeric Memory; TMT-B, Trail Making Test B (alphanumeric trail); Tower, Tower rearranging; VNR, Verbal Numeric Reasoning.

#### 2.2.4. Education and lifestyle data

Measures of education, alcohol consumption and smoking were derived from touchscreen questionnaire responses (Supplementary Materials 4). For education, a multiple-choice question allowed participants to report all qualifications. Participants were classified into one of five categories depending on the highest qualification they had attained. Alcohol consumption was measured as alcohol units per week (38). Past smokers were categorised by pack years (39) quartiles, with participants who had never regularly smoked in a fifth category. BMI (in kg/m^2^) was derived by the UK Biobank team from physical measurements.

### 2.3. Statistical Analysis

Statistical analyses were performed in R (version 3.2.3). Continuous variables were visually inspected and log-transformed where necessary to more closely approximate normal distribution. We reversed the TMT-B variable before statistical modelling, so that for all tests, higher scores represented better performance. For all outcome variables, negative regression coefficients thus indicated negative associations with cognitive performance. Regression coefficients are standardised throughout. Significance level was determined by a two-tail threshold of α = .05.

#### 2.3.1. Derivation of g-factor

We derived a *g*-factor as single measure of general cognitive functioning (40) by application of Principal Component Analysis (R function ‘prcomp’) including only complete observations (*n* = 13,589), extracting scores on the first principal component. This derived *g*-factor accounted for 45.0% of the test score variance. Component loadings of the model were all in the expected direction (loadings range 0.34 to 0.47; Supplementary Materials 5, Table S1 and Figure S3).

#### 2.3.2. Statistical Modelling

Associations between lifetime MDD and cognitive performance were tested using linear models. Each model included lifetime MDD as the predictor of interest and a cognitive score as the outcome variable. Primary models included age and sex as confounds (Tables 2 and 3). Additional models also included (i) education, (ii) lifestyle factors (alcohol, smoking and BMI), and (iii) both education and lifestyle factors (full model) as potential confounds. All results were false discovery rate (FDR) corrected across the 24 test-specific models and separately across the four *g*-factor models. Explorative analysis assessed age by MDD interactions on cognitive functioning.

**Table 2.** Demographic variables for participants who completed all cognitive tasks.

|  | *Control*<br>*n* = 5,221 | *Case*<br>*n* = 2,236 | *Effect Size* |
| --- | --- | --- | --- |
| **Age (years), mean (SD)<sup>a</sup>** | 64.7 (7.3) | 62.5 (7.1) | 0.30<sup>A***</sup> |
| **Sex female, *n* (%)<sup>b</sup>** | 2,379 (45.6) | 1,503 (67.2) | 0.20<sup>B***</sup> |
| **Education<sup>b</sup>** |  |  | n.s. |
| Incomplete, *n* (%) | 263 (5.0) | 87 (3.9) |  |
| Compulsory, *n* (%) | 600 (11.5) | 267 (11.9) |  |
| Continued, *n* (%) | 304 (5.8) | 121 (5.4) |  |
| College, *n* (%) | 1,327 (25.4) | 561 (25.1) |  |
| University, *n* (%) | 2,710 (51.9) | 1,188 (53.1) |  |
| Missing data, *n* (%) | 17 (0.3) | 12 (0.5) |  |
| **Alcohol units<sup>c</sup>** |  |  | 0.007<sup>C***</sup> |
| Median (IQR) | 10.7 (15.6) | 9.0 (14.9) |  |
| Missing data, *n* (%) | 317 (6.1) | 162 (7.2) |  |
| **Smoking** |  |  |  |
| Lifetime regular smoker, *n* (%)<sup>b</sup> | 1,079 (20.7) | 624 (27.9) | 0.08<sup>B***</sup> |
| Pack years, median (IQR)<sup>c</sup> | 14.8 (17.5) | 16.5 (22.4) | 0.005<sup>C**</sup> |
| Missing data, *n* (%) | 86 (1.6) | 34 (1.5) |  |
| **BMI<sup>c</sup>** |  |  | 0.006<sup>C***</sup> |
| Median (IQR) | 25.5 (5.0) | 26.2 (6.0) |  |
| Missing data, *n* (%) | 204 (4.6) | 112 (5.0) |  |

n.s. = non-significant , \* $P < .05$ , \*\* $P < .01$ , \*\*\* $P < .001$
<sup>a</sup> Independent t-test, <sup>A</sup> Hedge's $g$
<sup>b</sup> Chi-squared test, <sup>B</sup> Cramer's $V$
<sup>c</sup> Kruskal Wallis test, <sup>C</sup> $\eta^2$
BMI, Body Mass Index; IQR, Interquartile Range; SD, Standard Deviation.

**Table 3.** Linear model coefficients for the association between lifetime mood disorder and cognitive performance.

|  | **Model 1** | **Model 2** | **Model 3** | **Model 4** | * *P* |
| --- | --- | --- | --- | --- | --- |
|  | *Primary model* | *(+ Education)* | *(+ Lifestyle)* | *Full model* | < |
| ***g*-factor** |  |  |  |  | .05, |
| *β*-coefficient | **-0.10\*\*\*** | **-0.10\*\*\*** | **-0.08\*\*\*** | **-0.10\*\*\*** | ** |
| (95% CI) | (-0.15,-0.05) | (-0.15,-0.06) | (-0.14,-0.03) | (-0.15,-0.05) | *P* < |
| **DSST** |  |  |  |  | .01, |
| *β*-coefficient | **-0.13\*\*\*** | **-0.13\*\*\*** | **-0.12\*\*\*** | **-0.13\*\*\*** | *** |
| (95% CI) | (-0.18,-0.09) | (-0.18,-0.09) | (-0.17,-0.08) | (-0.18,-0.08) | *P* < |
| **TMT-B** |  |  |  |  | .00 |
| *β*-coefficient | **-0.08\*\*\*** | **-0.08\*\*\*** | **-0.08\*\*** | **-0.09\*\*\*** | 1, |
| (95% CI) | (-0.13,-0.04) | (-0.13,-0.04) | (-0.13,-0.03) | (-0.14,-0.04) | **bold** |
| **NM** |  |  |  |  | **d q** |
| *β*-coefficient | -0.03 | -0.03 | -0.02 | -0.03 | < |
| (95% CI) | (-0.08,0.02) | (-0.08,0.01) | (-0.07,0.03) | (-0.08,0.02) | .05 |
| **Matrix** |  |  |  |  | CI, |
| *β*-coefficient | -0.03 | -0.03 | 0.00 | -0.01 | Con |
| (95% CI) | (-0.07,0.02) | (-0.08,0.02) | (-0.05,0.06) | (-0.06,0.04) | fide |
| **VNR** |  |  |  |  | nce |
| *β*-coefficient | **-0.06\*\*\*** | **-0.06\*\*\*** | **-0.05\*** | **-0.07\*\*\*** | Inte |
| (95% CI) | (-0.10,-0.03) | (-0.10,-0.03) | (-0.09,-0.01) | (-0.10,-0.03) | rval |
| **Tower** |  |  |  |  | ; |
| *β*-coefficient | **-0.06\*** | **-0.06\*** | -0.05* | **-0.06\*** | DS |
| (95% CI) | (-0.11,-0.01) | (-0.11,-0.01) | (-0.11,0.00) | (-0.11,-0.01) | ST, |
|  |  |  |  |  | Dig |

it Symbol Substitution Task; *g*-factor, derived measure of general cognitive performance; Matrix, Matrix pattern completion; NM, Numeric Memory; TMT-B, Trail Making Test, alphanumeric trail; Tower, Tower rearranging; VNR, Verbal Numeric Reasoning.
\* $P < .05$ , \*\* $P < .01$ , \*\*\* $P < .001$
DSST, Digit Symbol Substitution Task; g-factor, derived measure of general cognitive performance; Matrix, Matrix pattern completion; NM, Numeric Memory; TMT-B, Trail Making Test, alphanumeric trail; Tower, Tower rearranging; VNR, Verbal Numeric Reasoning.

With the aim of investigating whether cognitive performance related to specific clinical features of depression, we also examined relationships, within the case group only, with recurrent depression, putative current depressive symptoms, impact on psychosocial functioning, and psychotropic medication use. Firstly, we tested a linear model including participants classified with lifetime MDD only (*n* = 2,179; Figure 2A) that predicted *g*-factor from all four clinical characteristics, including age and sex as covariates. Given the large degree of overlap between clinical characteristics within our sample, these variables were entered as simultaneous predictors in order to model their unique contributions to cognitive functioning. Secondly, we further explored whether the four clinical characteristics were associated with differential profiles of cognitive performance. Using the same approach, we tested another six univariate linear models (applying FDR-correction) that predicted each cognitive test scores from all four clinical characteristics, including age and sex as covariates.

**Figure 1.**
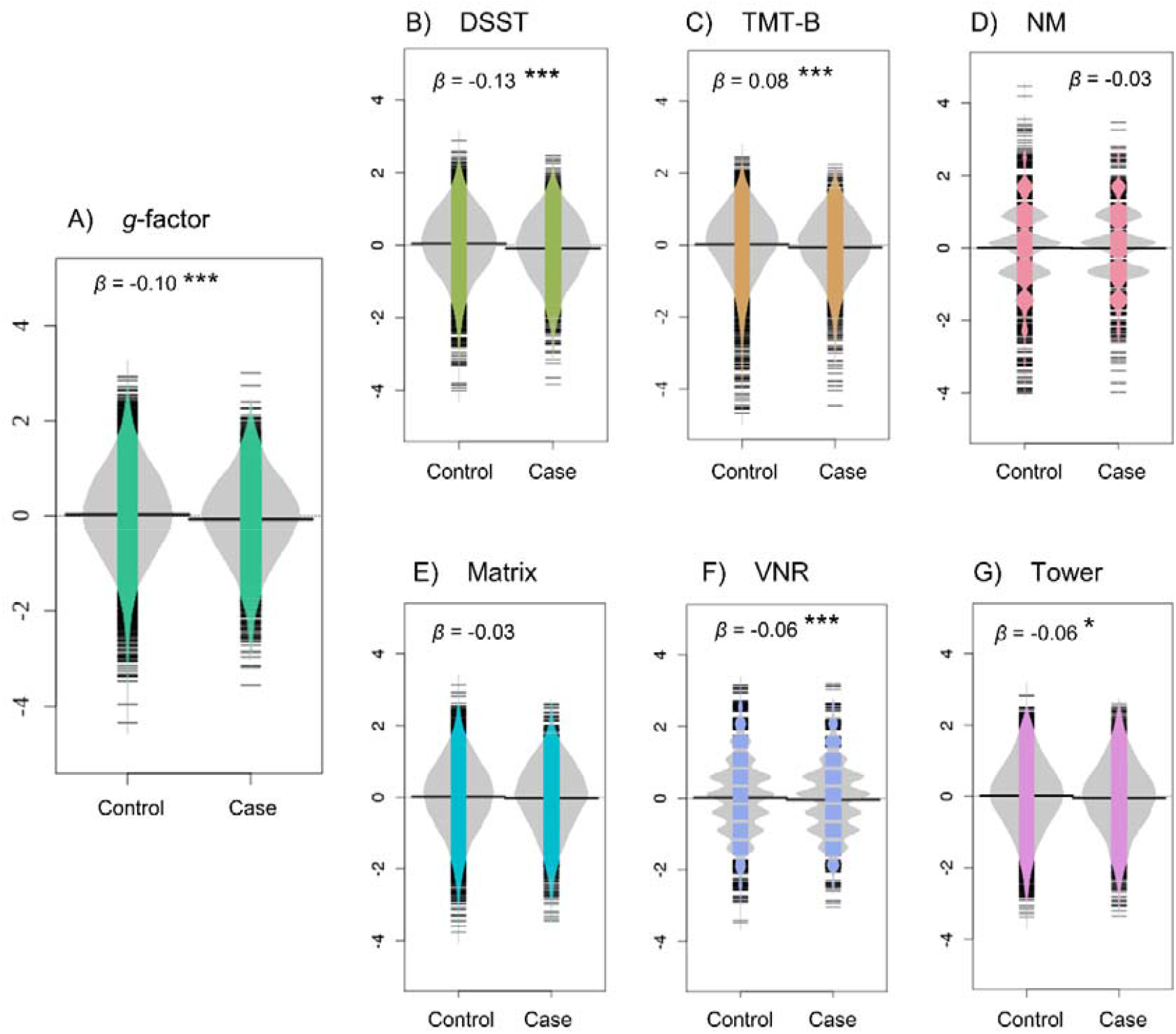
Visualisation of the base model results for (A) *g-*factor, (B) DSST, (C) TMT-B, (D) NM, (E) Matrix, (F) VNR, and (G) Tower. Graphs display case-control group differences in cognitive performance after adjustment for confounders (i.e. age and sex were regressed out). Specifically, they show significant but modest associations of lifetime MDD classification with lower general cognitive performance (*g*-factor), and with lower performance on DSST, TMT-B, VNR, and Tower. * *P* < .05, *\*\* P* < .01, *\*\*\* P* < .001 DSST, Digit Symbol Substitution Task; *g*-factor, derived measure of general cognitive performance; Matrix, Matrix pattern completion; NM, Numeric Memory; TMT-B, Trail Making Test, alphanumeric trail; Tower, Tower rearranging; VNR, Verbal Numeric Reasoning.

**Figure 2.**
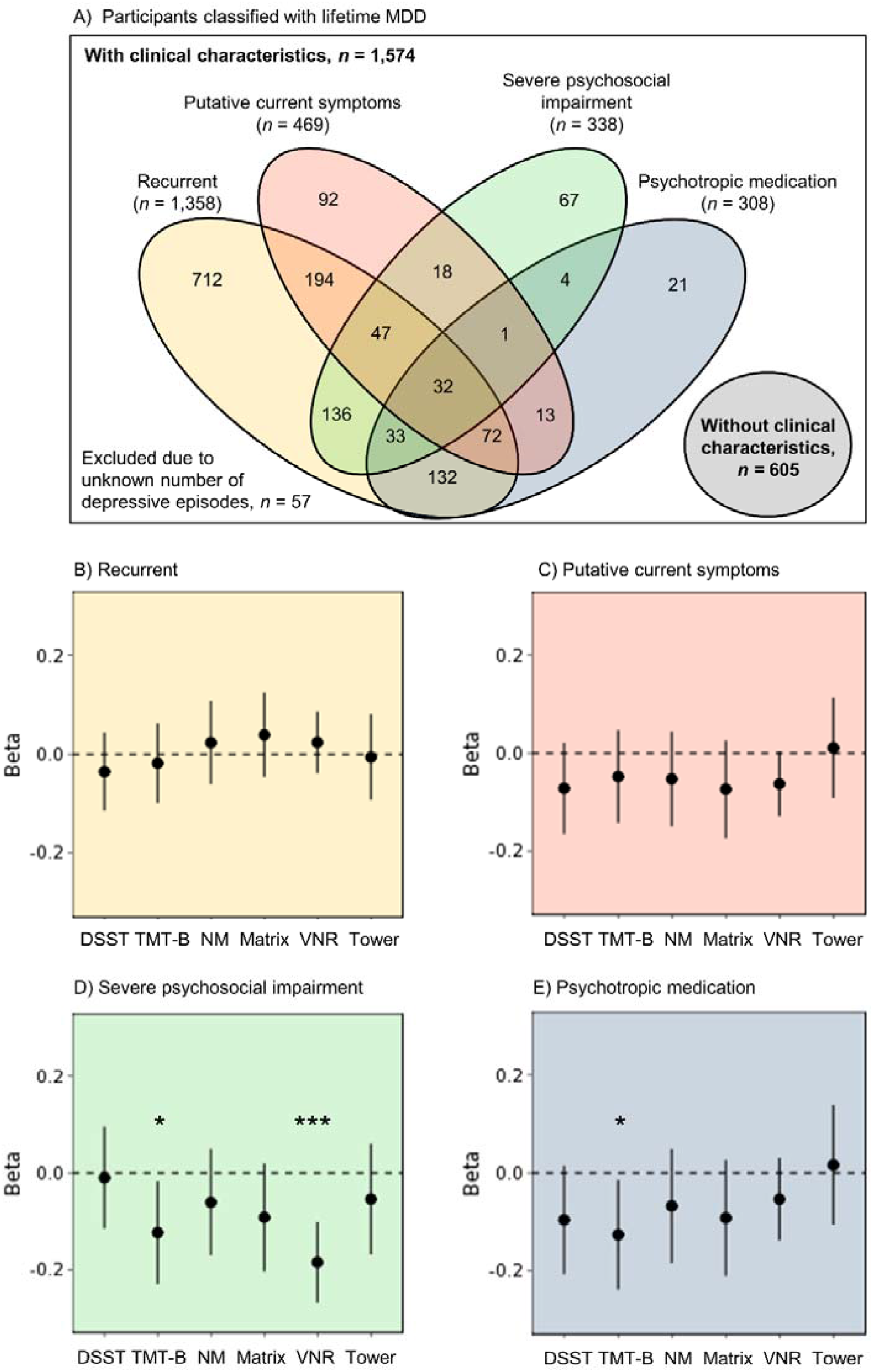
(A) Venn diagram of all participants classified with lifetime MDD and each of the four the clinical characteristics recurrent depression, putative current symptoms, severe psychosocial impairment, and psychotropic medication. These clinical characteristics were investigated within the group of case participants. (B-E) Cognitive profiles associated with the clinical characteristics (B) recurrent depression, (C) putative current symptoms, (D) severe psychosocial impairment (while symptomatic), and (E) use of psychotropic medication (at time of assessment). Points represent points estimates of the *β*-coefficient within the case models, whereas lines reflect the 95% confidence interval of the *β*-coefficient. * *P* < .05, *\*\* P* < .01, *\*\*\* P* < .001

## 3. Results

### 3.1. Sample Characteristics

Table 2 summarises demographic and lifestyle information on participants who completed all cognitive tests. The MDD group included a higher proportion of females (45.6% of controls, 67.2% of cases, χ^2^_(1)_ = 293.2, *P* < 2.2×10^−16^). MDD cases were also younger (*M* difference = 2.2 years, t_(4357.9)_ = 12.1, *P* < 2.2×10^−16^), had higher BMI (*Mdn* difference = 0.7 kg/m^2^, H_(1)_ = 40.1, *P* = 2.4×10^−10^), were more often lifetime regular smokers(20.7% of controls, 27.9% of cases, χ^2^_(1)_ = 46.0, *P* = 1.2×10^−11^), and among smokers, they had smoked more cigarettes (*Mdn* difference = 1.8 pack years, H_(1)_ = 7.8, *P* = 5.3×10^−3^). Conversely, control group participants reported more alcohol consumption (*Mdn* difference = 1.7 units/week, H_(1)_ = 46.6, *P* = 8.7×10^−12^). Education did not significantly differ between groups (χ^2^_(4)_ = 5.6, *P* =.23). Case and control sample sizes for individual cognitive tests, as well as test descriptive statistics, can be found in Table S2 (Supplementary Materials 6).

### 3.2. Associations between lifetime MDD and cognitive performance

Results indicated significant associations between lifetime MDD and cognitive performance (Table 3). The primary models consistently showed lower cognitive performance for the case group relative to controls with small effect sizes in terms of the *g*-factor (*β* = - 0.10, *P*_*FDR*_ = 4.7×10^−5^) and four of the six cognitive tests (DSST, *β* = −0.13, *P*_*FDR*_ = 1.1×10^−7^; TMT-B, *β* = −0.08, *P*_*FDR*_ = 1.1×10^−3^; VNR, *β* = −0.06, *P*_*FDR*_ = 2.0×10^−3^; Tower, *β* = −0.06, *P*_*FDR*_ = 3.4×10^−2^) (Figure 1). These results were robust over subsequent models that additionally included confounding variables, and almost all significant results (except for the Tower lifestyle model) survived multiple comparison correction. Associations between cognitive performance and covariates are reported in Supplementary Materials 7. Explorative analyses did not suggest any age by MDD interactions on cognitive functioning (Supplementary Materials 8, Table S4).

### 3.3. Associations between clinical characteristics and cognitive performance

For the associations between clinical characteristics and cognitive impairment, as measured by *g*-factor, we found no effect of recurrent depression (*β* = 0.02, *P* = .56), and a very modest, non-significant point estimate for putative current symptoms at time of assessment (*β* = −0.07, *P* = .15). Conversely, severe psychosocial impairment during the depressive episode was significantly associated with lower *g*-factor (*β* = −0.14, *P* = 1.5×10^−2^). Use of psychotropic medication showed a small, non-significant point estimate for its association with general cognitive functioning (*β* = −0.10, *P* = .10).

Further exploration within the lifetime MDD case group revealed differential cognitive profiles related to clinical characteristics (Figure 2B-E and Supplementary Materials 9, Table S5). Severe psychosocial impairment was associated with worse VNR performance (*β* = - 0.18, *P* = 1.2×10^−5^, *P*_*FDR*_ = 7.5×10^−5^). Furthermore, results indicated nominally significant associations with lower TMT-B performance for severe psychosocial impairment (*β* = −0.11, *P* = 2.3×10^−2^, *P*_*FDR*_ = 0.07) and psychotropic medication (*β* = −0.13, *P* = 2.7×10^−2^, *P*_*FDR*_ = 0.16).

## 4. Discussion

The findings of the current study describe a robust association between lifetime MDD and lower general cognitive performance within a population-based sample. The DSST, TMT-B, VNR and Tower results further indicate that executive functioning, processing speed and aspects of reasoning were predominantly affected. These effects were of modest overall effect size (*β* = −0.10 for general cognitive performance), suggesting that they are of limited clinical relevance, although such differences may have substantial consequences for whole populations. Comparisons within the case group however showed that severe impact on psychosocial functioning (while symptomatic) and use of psychotropic medication (at time of assessment) predicted the lowest measures of cognitive performance. These clinical characteristics also showed differential profiles of cognitive impairment, whereby severe psychosocial impairment was associated with reasoning (VNR) deficits. Severe psychosocial impairment, along with use of psychotropic medication, were also related to moderately lower mental flexibility and processing speed (TMT-B). These results highlight that cognitive functioning is impaired among individuals with current but also past MDD, and that deficits within specific cognitive domains may be more pronounced – and therefore of potential clinical relevance – in relation to impact of the depressive episode on psychosocial functioning (after recovery) and continued use of psychotropic medication.

Meta-analyses of small case-control studies have previously indicated cognitive deficits associated with current and remitted MDD within the domains of processing speed, executive functioning, memory and attention (4,12). This study corroborates such previous findings, but we also note that these population-level effect sizes are smaller than in traditional case-control studies. Of note, psychosocial impairment while symptomatic was also significantly associated with greater cognitive deficits in individuals with lifetime MDD, and we found some evidence for an association between psychotropic medication use and lower processing speed. This implicates that clinical studies that recruit from treatment centres, and thereby include participants from a patient group more likely to experience severe psychosocial impairment and/or use psychotropic medication, may show inflated effect sizes in comparison with the general population because of their sample characteristics.

Psychosocial impairment during the depressive episode was found to be associated to lower cognitive functioning subsequent assessment. Because other clinical characteristics such as the putative presence of current depressive symptoms were taken into account, this suggests that the impact of a depressive episode on quality of life may not be limited to the symptomatic phase. This result is consistent with previous adult clinical studies (23) and may reflect a subgroup of remitted individuals vulnerable to cognitive impairment, who potentially also experienced greater cognitive deficits during the depressive episode. Previous clinical research also showed that cognitive impairment in remitted MDD was associated with psychosocial dysfunction in multiple domains (24). Future research will need to address the underlying causality of the relationship between severe psychosocial impairment and impaired cognitive functioning in the context of lifetime MDD. If functional impairment persists specifically in remitted individuals who experience residual cognitive deficits, quality of life could be increased with interventions that target cognitive symptoms (24,41–43).

Findings of the current study did not indicate robust associations of cognitive impairment with recurrent depressive episodes current MDD symptoms, as well as less robust associations with psychotropic medication. Thus, within the healthy and non-clinical UK Biobank population, these MDD characteristics were less relevant to the association between lifetime MDD and cognitive impairment.

Of note, however, the current investigation was limited by the phenotypes available in UK Biobank, and one disadvantage was that no certain measure of remission or continuous measure of depressive symptoms at time of cognitive assessment could be established. Furthermore, although the current study benefited from the availability of newer, more reliable cognitive assessments in UK Biobank, this did impose the limitation of a cross-sectional design. We also note that the classification of lifetime MDD was based on the CIDI-SF, which, although based on diagnostic criteria and well validated, is still limited by reliance on retrospective self-report. Given the aim of the current study to investigate (residual) cognitive impairment associated with lifetime MDD in the general adult population, the mid-late life age range of UK Biobank participants at the time of the third assessment could also be considered a limitation. However, the cohort has been previously described as a relatively healthy mid-late life cohort (37), from which we excluded individuals with reported neurological or degenerative conditions from the current study, and exploration of age interaction effects did not suggest that results were driven by accelerated age-related cognitive decline. Future large-scale longitudinal research will be needed to unravel the relationship between MDD and cognitive impairment over the disease course as well as the lifespan.

In summary, the present findings suggest that lifetime MDD relates to impaired cognitive functioning among adults in their mid-late life, with most prominent deficits in the cognitive domain of processing speed (DSST). Severe psychosocial impairment during the depressive episode was associated with greater overall cognitive impairment, and specifically on tasks of reasoning (VNR) and mental flexibility and processing speed (TMT-B). Furthermore, clinical characteristics showed differential profiles of impairment of modest effect. These findings add to meta-analytic evidence by providing accurate population-level estimates, which is an important foundation for future studies addressing cognitive functioning in the context of MDD. Longitudinal trajectories of cognitive performance in lifetime MDD and the differential influences of pharmacological treatments on these trajectories are important targets for further research. The longitudinal association between psychosocial impairment while symptomatic and subsequent cognitive impairment, also when remitted, reflects their likely impact on quality of life and suggests that both cognitive and psychosocial functioning should be key targets in the treatment of MDD.

## Data Availability

This study used data from UK Biobank, a resource that is available upon registration. The dataset in the form used for the current study may be shared upon request.

## Supporting information

Appendices can be found in the online version of this article. The R code of our statistical analysis will be made available via [link] after publication.

## Acknowledgements

This research has been conducted using the UK Biobank resource. We would like to thank all of the participants from UK Biobank who have shown their continued commitment, as well as the UK Biobank team for collecting and providing the data.

